# Serum Neurofilament Light Chain and Glial Fibrillary Acidic Protein in Multiple Sclerosis: A Disease-Stage Gradient from Relapsing to Progressive Disease on a Commercial ECLIA Platform (n=603)

**DOI:** 10.64898/2026.06.24.26356462

**Authors:** Nicholas S. Streicher

## Abstract

**Background:** Serum neurofilament light chain (NfL) indexes axonal injury and glial fibrillary acidic protein (GFAP) astrocytic pathology in multiple sclerosis (MS). GFAP rises disproportionately as relapsing-remitting MS (RRMS) shifts to progressive forms on research-grade SIMOA. The commercial Roche Elecsys ECLIA platform reads six-fold lower and is undescribed across subtypes.

**Objective:** To describe both markers by MS subtype on ECLIA.

**Methods:** Retrospective single-center analysis of 603 MS patients (2022–2026). NfL and GFAP were measured by LabCorp Roche Elecsys ECLIA; subtype came from ICD-10 codes and notes. We examined both markers by subtype, their correlation, and NfL against gadolinium-enhancing (Gd+) MRI lesions.

**Results:** Median NfL was 1.32 pg/mL (IQR 1.01–1.91). Both rose with stage, steeper for GFAP: NfL 1.18 (RRMS), 1.54 (SPMS, p<0.001), 1.78 (PPMS, p=0.001); GFAP 41.90, 63.80 (p<0.0001), 75.75 (p=0.08, n=6). SPMS and PPMS GFAP did not differ (p=0.83). The markers correlated moderately (r=0.569). Of 34 Gd+ encounters with NfL within 30 days, 3 (9%) were elevated.

**Conclusion:** On ECLIA, both markers rose with MS stage, GFAP more steeply, and both progressive subtypes exceeded RRMS. NfL rarely flagged a recent Gd+ lesion, consistent with its delayed kinetics. The two index distinct processes and reproduce on an orderable assay a profile once confined to research-grade SIMOA.

## INTRODUCTION

Serum NfL is the most validated blood biomarker of MS disease activity, supported by multinational consortium guidance^1^ and individual prognostication frameworks^2,3^. It is a non-specific marker of neuro-axonal injury^4^ whose blood levels rise and resolve over months around an injury^5^; circulating levels therefore reflect the rate of axonal loss integrated over recent weeks to months rather than any single event.

NfL behaves as a trajectory biomarker rather than a lesion detector. In the RESTORE natalizumab-interruption study, 71% of participants with Gd+ lesions never crossed the 95th-percentile serum NfL threshold despite confirmed focal inflammation, and the rise that did occur peaked a median of ∼8 weeks after the lesion appeared**—**delayed and modest, even with monthly SIMOA sampling^6^. In patients on high-efficacy therapy, breakthrough lesions are small and the resulting axonal signal is modest against background turnover. The clinical value of NfL lies in identifying patients whose cumulative rate of axonal loss exceeds expectation^7,8^.

This rate-versus-burden split has a CNS-specific basis. NfL is released by injured axons and clears within weeks, so when the injurious process quiets, serum NfL falls toward normal—a low value signals quiet disease, not repair, because CNS axons largely do not regenerate, unlike peripheral nerve. The damage already done persists and is captured instead by accumulated astrogliosis (GFAP) and atrophy. A patient can therefore carry a normal NfL with a high GFAP—current injury controlled, cumulative burden already laid down—which is why a single marker can mislead and the two are not interchangeable.

GFAP, an astrocytic protein, captures pathology that NfL does not. Across cohorts GFAP rises disproportionately to NfL as disease shifts from relapsing to progressive^9,10,11,12^ and predicts progression independent of relapse activity^13,14^; under disease-modifying therapy, GFAP and NfL track different mechanisms^15,16^. Pairing the two markers is a clinically accessible step beyond either alone.

Most of this evidence derives from the research-grade SIMOA platform, which has limited clinical availability. The commercially orderable Roche Elecsys ECLIA platform yields values approximately 85% lower than SIMOA (r=0.991)^17^ and is not interchangeable with it^18^. Karam validated ECLIA NfL in peripheral neuropathy but did not include MS^19^. We therefore characterized serum NfL and GFAP on the commercial ECLIA platform in a large real-world MS cohort. Our objectives were to describe both markers across all three MS subtypes, to quantify their correlation, and to assess NfL–MRI concordance. We hypothesized that both markers would rise from relapsing to progressive disease—more steeply for GFAP—that they would correlate only moderately, indicating complementary information, and that NfL would seldom be elevated on samples drawn near enhancing lesions, given its delayed, integrated kinetics.

## METHODS

### Study Design

Retrospective single-center analysis of patients with multiple sclerosis (MS) seen at Georgetown University Hospital between January 2022 and March 2026, drawing on structured and free-text electronic health record (EHR) data. The protocol was IRB-approved (protocol no. STUDY00008842) with waiver of informed consent.

### Inclusion and Exclusion Criteria

Patients were eligible if they carried an MS diagnosis (ICD-10 G35 or a granular G35.A/B/C code) and had at least one serum NfL or GFAP result on the LabCorp Roche Elecsys ECLIA platform during the study window. Patients without an ECLIA biomarker result were excluded. No restriction was placed on age, sex, treatment status, or disease duration.

### Participant Characteristics

Demographic and subtype composition are summarized in Table 1. The cohort was predominantly female with a mean age near 50 years. A documented subtype was available for a minority of patients—RRMS most common, then SPMS and PPMS—while the remainder carried only the generic G35 code and were classified MS-unspecified.

**Table 1.**
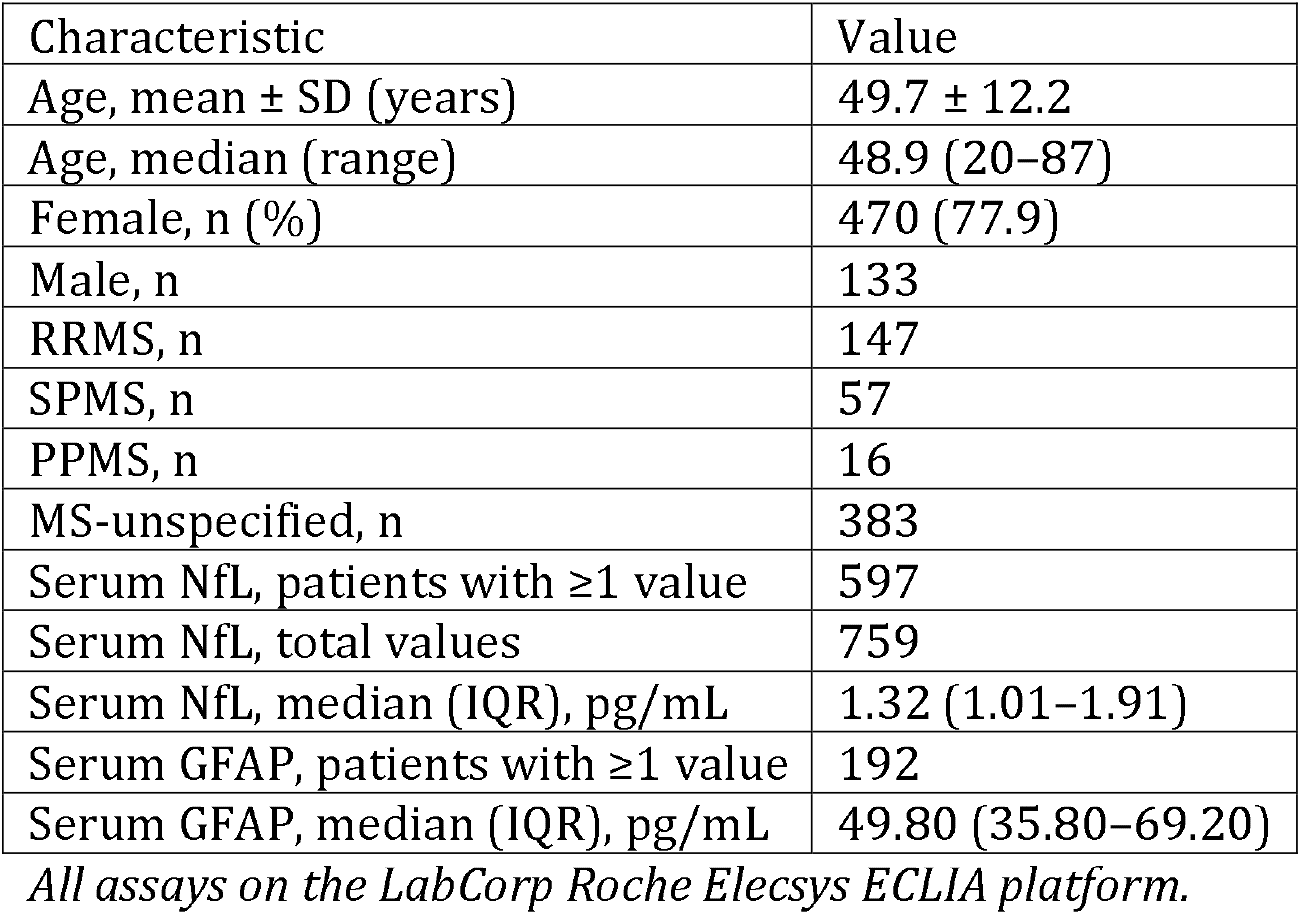
Cohort characteristics and first-per-patient serum biomarker values.

| Characteristic | Value |
| --- | --- |
| Age, mean ± SD (years) | 49.7 ± 12.2 |
| Age, median (range) | 48.9 (20–87) |
| Female, n (%) | 470 (77.9) |
| Male, n | 133 |
| RRMS, n | 147 |
| SPMS, n | 57 |
| PPMS, n | 16 |
| MS-unspecified, n | 383 |
| Serum NfL, patients with $\geq 1$ value | 597 |
| Serum NfL, total values | 759 |
| Serum NfL, median (IQR), pg/mL | 1.32 (1.01–1.91) |
| Serum GFAP, patients with $\geq 1$ value | 192 |
| Serum GFAP, median (IQR), pg/mL | 49.80 (35.80–69.20) |

*All assays on the LabCorp Roche Elecsys ECLIA platform.*

### Sampling Procedures

Eligible patients were identified by querying the EHR for MS ICD-10 codes across the study window; all who met criteria were analyzed, without random selection (a consecutive, convenience sample). MS subtype was classified primarily from granular ICD-10 codes (G35.A RRMS, G35.B PPMS, G35.C SPMS), with clinical-note terminology as fallback; where codes conflicted, the more progressive subtype was retained, and patients with only the legacy generic code were classified MS-unspecified. Subtype was never inferred from age or biomarker values; where both a granular code and note terminology were available, the two agreed in 97% of patients. Both serum and plasma GFAP results were retained as a single biomarker stream, consistent with routine ordering, and repeat draws on the same calendar date were collapsed to one timepoint. MRI reports were deduplicated to one record per encounter.

### Sample Size, Power, and Precision

Sample size was set by data availability rather than an a priori power calculation; all eligible patients in the window were analyzed. The cohort comprised 603 patients, of whom 186 contributed paired same-day NfL and GFAP and 396 contributed 454 MRI encounters. Subgroup precision varied: RRMS and SPMS were well represented, whereas PPMS was small—particularly for GFAP—so PPMS estimates are imprecise and interpreted cautiously. Analyses are descriptive, with no correction for multiple comparisons.

### Measures and Covariates

Serum NfL, GFAP, and S-100B were measured by LabCorp Roche Elecsys ECLIA. Booth reported an 85.1% negative bias versus research-grade SIMOA with high correlation (r=0.991; SIMOA = 6.566 × ECLIA − 0.812)^17^; inter-assay comparisons confirm the platforms are strongly correlated^20^ but not interchangeable^18^. MRI measures included new T2/FLAIR lesions, gadolinium enhancement (present or absent), T2 lesion burden, stability, brain and cord atrophy, and lesion distribution; patient-level MRI activity was classified as documented active enhancement, documented stable, mixed history, or no MRI language available. Age and MS subtype were the prespecified covariates.

### NfL–MRI Matching

NfL–MRI analyses used NfL values drawn within a 30-day window of each MRI encounter, with subgroup analyses tightened to ≤7 days. This narrow window was chosen to associate NfL with recent MRI activity and to limit confounding from new lesions, uncertain lesion timing, and medication changes that grows with longer intervals. As a sensitivity analysis, the post-MRI window was extended to 120 days, past the ∼8-week median time to peak serum NfL reported by Fox^6^. For the lesion-burden analysis (Figure 2), NfL drawn near a documented enhancing lesion was expressed as percent deviation from an age-expected baseline derived by log-linear regression of NfL on age in patients without documented enhancement; ten patients with both a quiescent (>120-day) and a near-lesion (<60-day) draw provided a within-patient change.

### Statistical Analysis

NfL–GFAP correlation was assessed by Spearman rank correlation in patients with paired same-day measurements (n=186, first per patient). Subtype comparisons used the Mann–Whitney test, and within-subtype age effects on each marker were assessed by Spearman correlation. Continuous variables are reported as median (interquartile range) unless otherwise stated.

## RESULTS

### Cohort

The cohort comprised 603 MS patients (mean age 49.7 ± 12.2 years; median 48.9, range 20–87; 470 female, 77.9%). Subtype was documented for 220 patients (36%): 147 RRMS, 57 SPMS, and 16 PPMS; the remaining 383 were MS-unspecified (Table 1).

### Serum NfL Distribution

There were 759 NfL values from 597 patients; the first-per-patient median was 1.32 pg/mL (IQR 1.01–1.91, range 0.42–25.80). Sixty-four patients (10.7%) had a first value flagged elevated against the assay’s age-specific reference interval^17^. Longitudinal NfL sampling (≥2 timepoints) was available for 133 patients.

### NfL and GFAP by MS Subtype

Both biomarkers showed a monotonic disease-stage gradient, with GFAP rising more steeply. Median NfL was 1.18 pg/mL in RRMS, 1.54 in SPMS (+31% vs RRMS, p<0.001), and 1.78 in PPMS (+51% vs RRMS, p=0.001). Median GFAP was 41.90 pg/mL in RRMS, 63.80 in SPMS (+52% vs RRMS, p<0.0001), and 75.75 in PPMS (+81% vs RRMS, p=0.08, n=6 underpowered). SPMS and PPMS GFAP did not differ (p=0.83) (Table 2, Figure 1).

**Table 2.**
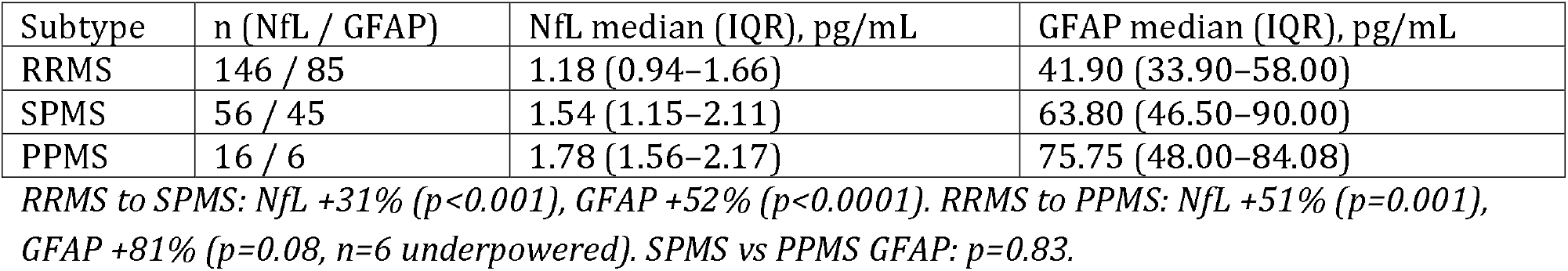
Serum NfL and GFAP by MS subtype (first-per-patient medians).

**Figure 1.**
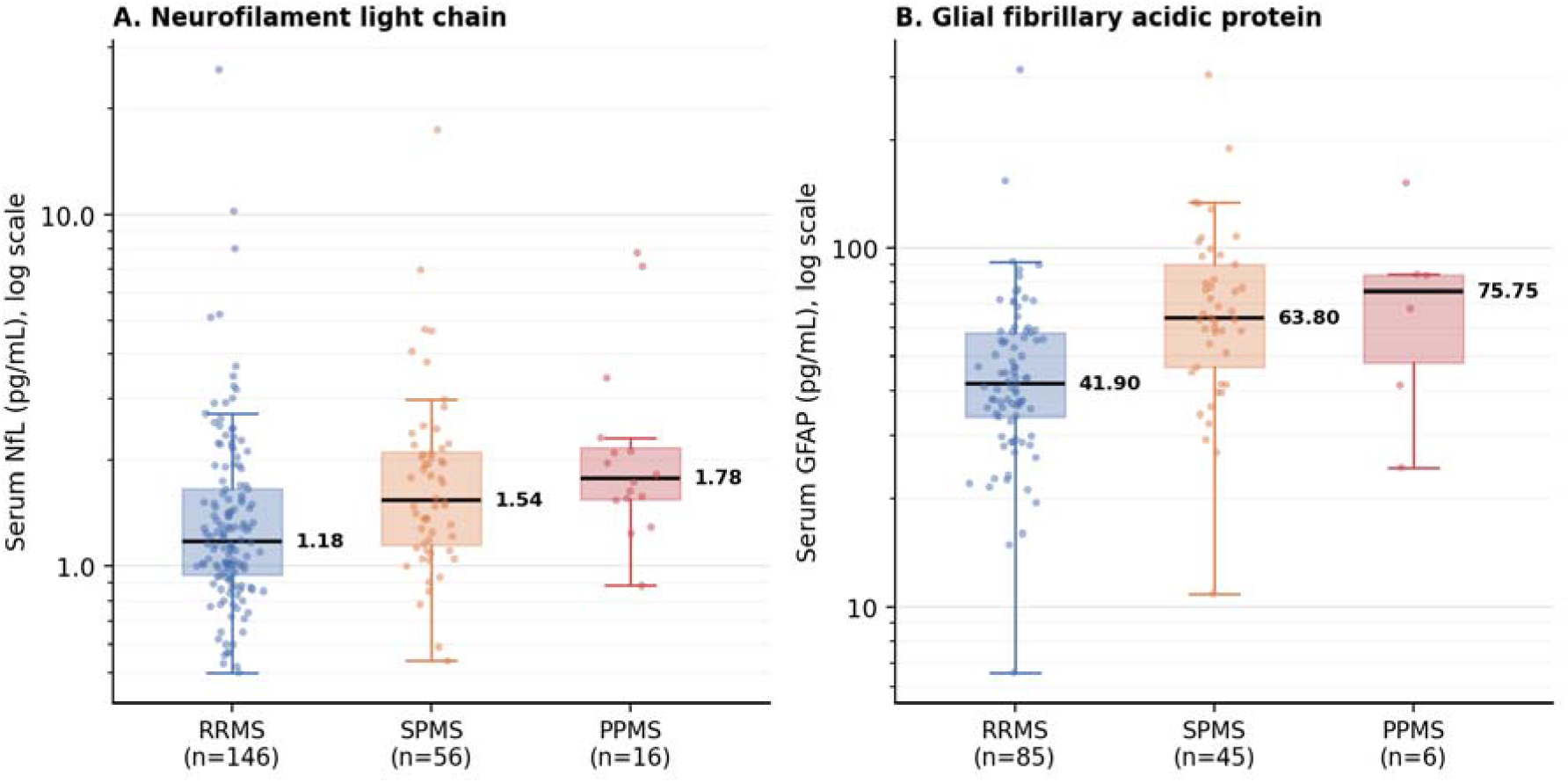
Serum NfL (A) and GFAP (B) by MS subtype (first-per-patient values); box plots with individual points on a log scale, medians and n shown. See Results.

### NfL–GFAP Correlation

In 186 patients with paired same-day measurements, NfL and GFAP were moderately correlated (Spearman r=0.569, p<0.001), leaving most of each marker’s variance independent of the other. Relative to RRMS, SPMS patients ranked higher on GFAP than on NfL (GFAP-dominant; p=0.037); age and clinical-note features did not separate NfL-dominant from GFAP-dominant patients.

### GFAP and Age

GFAP correlated with age across the cohort (Spearman r=0.51, n=192, p<0.0001). Within-subtype correlations were also significant: RRMS r=+0.38 (p<0.001, n=85), SPMS r=+0.33 (p=0.027, n=45); PPMS (n=6) was too small to test. NfL showed similar patterns (RRMS r=+0.45, p<0.001; SPMS r=+0.52, p<0.001), consistent with published age-specific NfL reference intervals^17^. Age and disease stage both contribute to GFAP variation rather than one explaining the other away.

### NfL and MRI Disease Activity

Of 454 MRI encounters across 396 of 603 patients, 42 carried documentation of active gadolinium enhancement. Thirty-four of these had NfL drawn within 30 days (median 1.21 pg/mL); 3 (9%) showed elevated NfL (>2.5 pg/mL). Restricting to NfL drawn within 7 days (n=20) yielded a median of 1.24 pg/mL, indistinguishable from the 30-day result (Supplementary Table S1). Extending the post-MRI window toward the reported ∼8-week peak surfaced one further elevation, at 42 days (3.75 pg/mL) near an enhancing pontine lesion; no other late draw was elevated. Across all 41 patients with NfL near a documented enhancing lesion, values clustered at low lesion number and near an age-expected baseline; four exceeded the elevation threshold, and none reached the high lesion burden at which Fox’s median rise crosses it (Figure 2).

**Figure 2.**
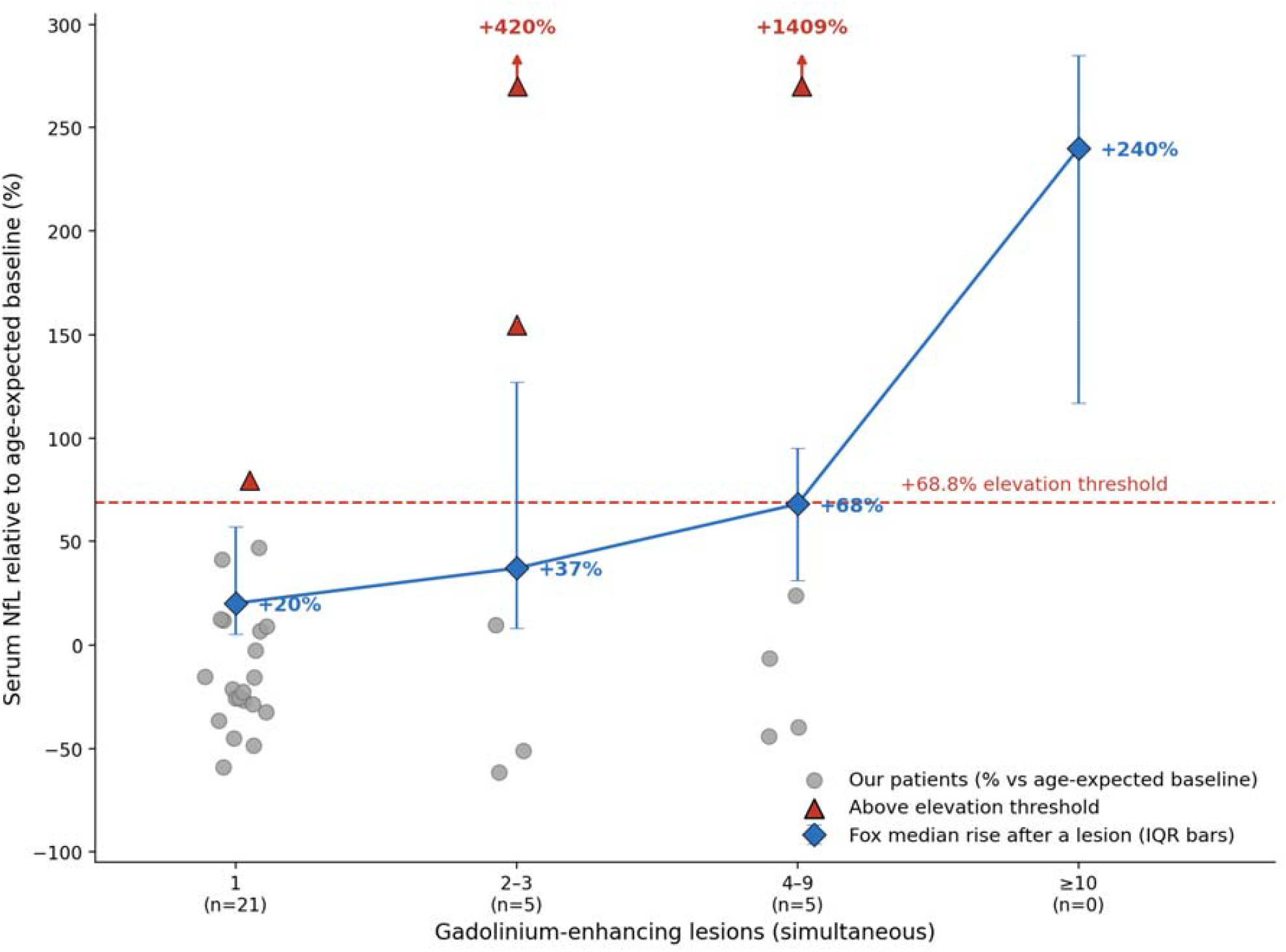
Serum NfL near gadolinium-enhancing lesions by number of simultaneous lesions. Patients sampled near a documented enhancing lesion (n=41; the 31 with a documented simultaneous-lesion count are plotted), as percent deviation from an age-expected baseline; gray circles are individual patients and red triangles those exceeding the elevation threshold (+68.8%, Fox/RESTORE’s 95th percentile; two off-scale, arrows). Blue diamonds: published peak median rise by lesion number (Fox/RESTORE^6^), interquartile bars. See Results and Discussion.

## DISCUSSION

### Principal Findings

On the commercially orderable ECLIA platform, this cohort reproduced the dual-biomarker architecture previously characterized mainly on SIMOA^9^. Both markers rose with MS stage, GFAP more steeply, so both progressive subtypes stood above RRMS and GFAP in PPMS matched SPMS. The two markers correlated only moderately, leaving most of each one’s variance independent. NfL was rarely elevated on samples drawn near an enhancing lesion. These findings address the study’s three aims—the dual-biomarker profile across all three subtypes, the NfL–GFAP correlation, and NfL–MRI concordance—on a commercially available assay.

### Biomarker Profiles Across MS Subtypes

NfL reflects the current rate of axonal injury^3,4^—rising with activity, falling when treatment suppresses it^8,21^. GFAP reflects accumulated astrocytic activation that persists^9,11^. Both rose across RRMS, SPMS, and PPMS, GFAP more steeply, and the gradient tracks what drives each stage. Relapsing disease injures axons through acute inflammatory lesions, producing transient NfL release^5,6^. Progressive disease shifts to chronic astrocytic pathology, where axonal loss is slow and diffuse but astrocytic burden accrues^13,14^. The disproportionate GFAP rise is the serum signature of progression^9,10,12^. The relapsing-to-progressive distinction likely reflects an underlying pathological shift that the clinical course and the serum markers track in parallel.

PPMS progresses from onset without a relapsing phase, so its accumulated astrocytic burden should match or exceed that of SPMS, where the progressive phase is shorter on average. That is what we observe: GFAP in PPMS sits at or above the SPMS level, and the two subtypes are statistically indistinguishable. Both appear to converge on a similar astrocytic burden once progression is established^9,12^, whatever route brought them there. The small PPMS subgroup makes this suggestive rather than definitive, but it follows the prediction of an accumulation model.

### Comparison With Published Data

These patterns reproduce, on a commercial ECLIA platform, what research-grade SIMOA established: a GFAP rise that follows disease type and severity^11,12^, tracks progression independent of relapse activity^9,10,13,14^, and persists under therapy^15,16^. Absolute concentrations are far lower here^17,18^, so the reproducible finding is direction, not magnitude. Our proportion of patients with NfL elevated for age falls between published cohorts—below a larger real-world clinic population^22^ and above tightly controlled trials—a selection gradient from practice through clinic to trial.

Earlier subtype-resolved serum work was mostly two-group and SIMOA-based; Ayrignac found the two markers correlated yet only GFAP separated PPMS from RRMS^12^. We extend this to the full triad on an orderable assay, same hierarchy, steeper GFAP gradient. The markers are only moderately correlated, so most variance is independent—the basis for measuring both. SPMS leans toward GFAP-dominance, echoing the higher GFAP-to-NfL ratio reported in progressive disease^13,14^. Whether GFAP predicts progression when NfL is low, our cross-sectional data cannot test.

GFAP tracks accumulated disease rather than current activity^13,14^. GFAP did not differ between patients with and without documented enhancement, and those with stable MRI carried slightly higher levels—indicating advanced or closely followed disease, not acute inflammation. GFAP also rose with age within subtypes, and the cohort-wide effect partly reflects the subtype gradient, since progressive patients are older; age-specific reference intervals are therefore essential^17^. Age is not a confounder to remove but a second axis along which astrocytic burden accumulates; aging and progression are both time-integrated exposures that the marker sums.

### Serum Biomarkers and MRI

NfL rarely flagged individual enhancing lesions. One lesion raises NfL only modestly above a patient’s own baseline—a real within-person change, too small to clear a population or age-referenced threshold, so a single spot value reads normal; the rise also scales with lesion number and peaks weeks later^6^. Our spot draws, referenced to an age baseline with no per-patient pre-lesion value, sat at that baseline (Figure 2). The marker is thus specific but not sensitive—a raised value rules activity in, a normal one cannot rule it out, like a troponin drawn too early in chest pain—so its utility depends on when it is checked, which differs across diseases by tempo.

Lesion location also matters. The spinal cord packs long tracts densely, so a cord lesion severs more long-tract axons than a comparable cerebral plaque; serum NfL rises stepwise from no enhancement, to brain or cord enhancement, to both^3^. The source need not be central at all: MR neurography reveals peripheral nerve involvement in MS itself—T2-hyperintense nerve lesions in relapsing patients^23^. Routine MRI images only the brain, so cord and peripheral-nerve activity driving NfL can go unseen—one reason a raised marker may accompany a quiet brain scan.

### Clinical Implications

MRI and the serum markers carry complementary information. MRI localizes focal lesions; the markers index diffuse processes imaging does not capture directly—NfL the current rate of axonal loss^3,7,8^, GFAP the accumulated burden of progression^9,14^. Serum markers cannot localize a lesion, and imaging cannot quantify the tempo or burden they report. Read together they are more informative than either alone; they add to imaging rather than replace it.

The two-marker pattern carries distinct information. A raised NfL points to active axonal injury and tracks treatment response^21^, whereas a GFAP-dominant pattern—GFAP elevated with NfL near baseline—points to progression that focal MRI may miss. The same axis separates MS from AQP4-positive neuromyelitis optica spectrum disorder, an astrocytopathy at the GFAP-dominant extreme^24^. Concordant elevation of both markers, by contrast, indicates active inflammation superimposed on accumulating progression^13,14^—the profile of active progressive disease.

### Limitations

This was a single-center retrospective study. Subtype was undocumented for most of the cohort, limiting subtype comparisons—PPMS especially, where GFAP rested on only a few patients; therapy subgroups and EDSS were unavailable. MRI findings were extracted categorically from clinical notes; a single Gd+ scan dates enhancement, not lesion onset, so without serial imaging the lesion-to-NfL interval and its delayed peak were untestable. Because care came from one health system, scans and laboratory draws obtained elsewhere were unavailable, leaving NfL–MRI pairs incomplete. Serial draws were too sparse to resolve subtype trajectories (Supplementary Figure S1). Age-specific ECLIA GFAP reference intervals do not yet exist^17^, so interpretation rests on cohort-level patterns rather than individual cutoffs.

## CONCLUSIONS

NfL and GFAP are integrative markers, not snapshots of acute injury. Like HbA1c, each sums damage over a characteristic window—NfL over weeks to months, GFAP over years—rising only when injury is large in aggregate: sustained, repeated, or a single severe insult such as many simultaneous lesions. Read as a pair against a patient’s own baseline^21^, they resolve the two processes that define MS course—active axonal injury and accumulated progression—that neither a single marker nor a scan captures alone.

The advance is access. A dual-marker profile once confined to research-grade SIMOA laboratories now reproduces on an assay any clinician can order, bringing serial neuro-axonal and astrocytic monitoring within reach of routine MS care. What a cross-sectional snapshot cannot yet show— whether these profiles forecast an individual’s trajectory—now needs prospective, longitudinally sampled cohorts with peak-timed draws, outcome linkage, and age-specific ECLIA reference intervals^17^.

## Acknowledgements

None.

## Declaration of Conflicting Interests

The author declares no potential conflicts of interest with respect to the research, authorship, and/or publication of this article.

## Funding

This research received no specific grant from any funding agency in the public, commercial, or not-for-profit sectors.

## Ethics Approval and Informed Consent

This retrospective study was approved by the Georgetown University Institutional Review Board (protocol no. STUDY00008842; initially approved October 10, 2025), with a waiver of informed consent for review of existing electronic health record data.

## Data Availability Statement

The data that support the findings of this study are not publicly available because they contain information that could compromise the privacy of research participants. De-identified data may be available from the author on reasonable request, subject to institutional approval.

## Author Contributions

The sole author (N.S.) conceived and designed the study, performed the data extraction and statistical analysis, interpreted the results, and drafted and revised the manuscript.

## Use of Artificial Intelligence

The author used a large language model (Anthropic Claude) to assist with copy editing and manuscript preparation. The author reviewed and verified all content, data, and citations, and takes full responsibility for the integrity of the work.

## Notes

### Competing Interest Statement

The authors have declared no competing interest.

### Author Declarations

The Institutional Review Board of Georgetown University waived ethical approval for this work (protocol no. STUDY00008842), with a waiver of informed consent for review of existing electronic health record data.

