## Supplemental Material for "Serum Neurofilament Light Chain and Glial Fibrillary Acidic Protein in Multiple Sclerosis: A Disease-Stage Gradient from Relapsing to Progressive Disease on a Commercial ECLIA Platform (n=603)"

### Supplementary Material

**Supplementary Table S1. Serum NfL in gadolinium-enhancing MRI encounters.**

| Window | n | Median NfL, pg/mL | Elevated (>2.5 pg/mL), n (%) |
| --- | --- | --- | --- |
| Within 30 days | 34 | 1.21 | 3 (9) |
| Within 7 days | 20 | 1.24 | — |

*NfL drawn within the stated window of a documented gadolinium-enhancing MRI encounter. See main-text Discussion.*

**Supplementary Figure S1. Within-patient biomarker trajectories by MS subtype.** Patients with ≥2 draws ≥30 days apart (faint), normalized to first value (=50); subtype averages (bold). (A) NfL (n=132), trajectory direction not differing by subtype (progressive vs RRMS p=0.78); (B) GFAP, RRMS only (n=7). See Discussion.

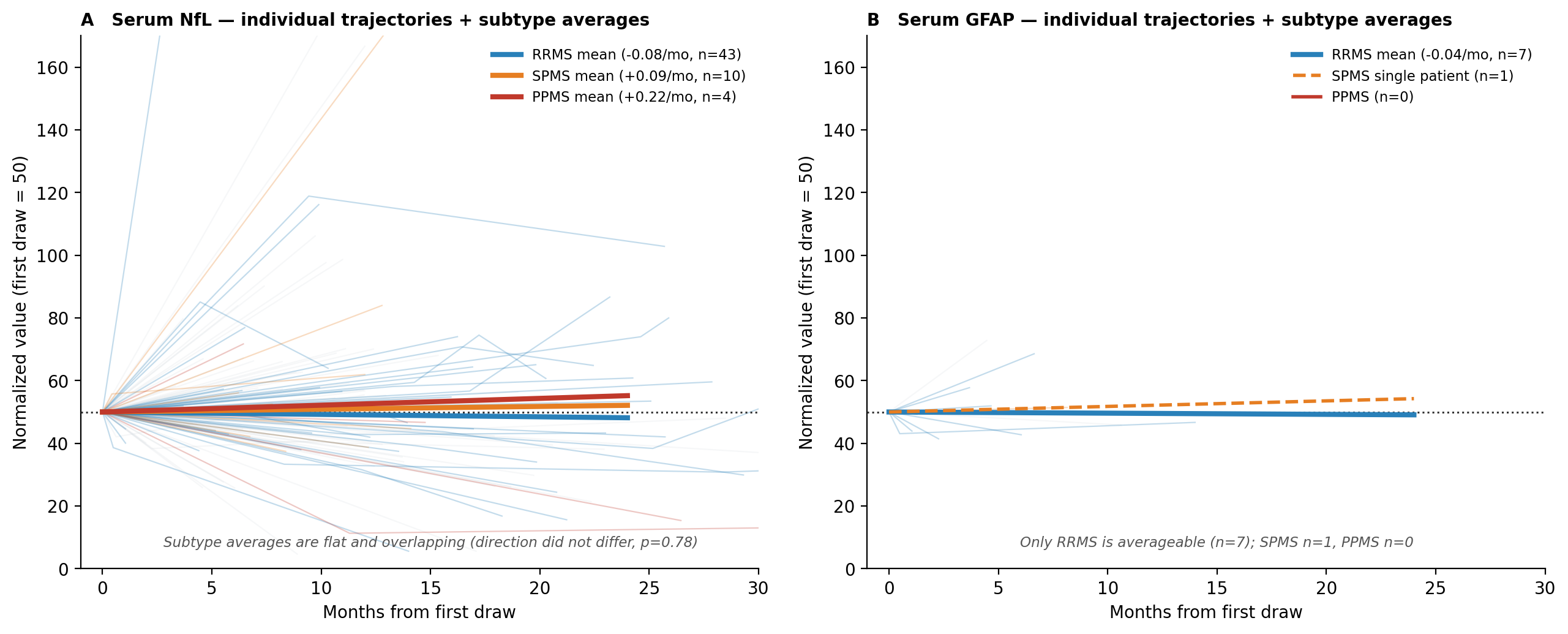
